# Spatial Dissection of the Distinct Cellular Responses to Normal Aging and Alzheimer’s Disease in Human Prefrontal Cortex at Single-Nucleus Resolution

**DOI:** 10.1101/2024.05.21.24306783

**Authors:** Yun Gong, Mohammad Haeri, Xiao Zhang, Yisu Li, Anqi Liu, Di Wu, Qilei Zhang, S. Michal Jazwinski, Xiang Zhou, Xiaoying Wang, Lindong Jiang, Yi-Ping Chen, Xiaoxin Yan, Russell H. Swerdlow, Hui Shen, Hong-Wen Deng

## Abstract

Aging significantly elevates the risk for Alzheimer’s disease (AD), contributing to the accumulation of AD pathologies, such as amyloid-β (Aβ), inflammation, and oxidative stress. The human prefrontal cortex (PFC) is highly vulnerable to the impacts of both aging and AD. Unveiling and understanding the molecular alterations in PFC associated with normal aging (NA) and AD is essential for elucidating the mechanisms of AD progression and developing novel therapeutics for this devastating disease. In this study, for the first time, we employed a cutting-edge spatial transcriptome platform, STOmics® SpaTial Enhanced Resolution Omics-sequencing (Stereo-seq), to generate the first comprehensive, subcellular resolution spatial transcriptome atlas of the human PFC from six AD cases at various neuropathological stages and six age, sex, and ethnicity matched controls. Our analyses revealed distinct transcriptional alterations across six neocortex layers, highlighted the AD-associated disruptions in laminar architecture, and identified changes in layer-to-layer interactions as AD progresses. Further, throughout the progression from NA to various stages of AD, we discovered specific genes that were significantly upregulated in neurons experiencing high stress and in nearby non-neuronal cells, compared to cells distant from the source of stress. Notably, the cell-cell interactions between the neurons under the high stress and adjacent glial cells that promote Aβ clearance and neuroprotection were diminished in AD in response to stressors compared to NA. Through cell-type specific gene co-expression analysis, we identified three modules in excitatory and inhibitory neurons associated with neuronal protection, protein dephosphorylation, and negative regulation of Aβ plaque formation. These modules negatively correlated with AD progression, indicating a reduced capacity for toxic substance clearance in AD subject samples. Moreover, we have discovered a novel transcription factor, ZNF460, that regulates all three modules, establishing it as a potential new therapeutic target for AD. Overall, utilizing the latest spatial transcriptome platform, our study developed the first transcriptome-wide atlas with subcellular resolution for assessing the molecular alterations in the human PFC due to AD. This atlas sheds light on the potential mechanisms underlying the progression from NA to AD.

## INTRODUCTION

Alzheimer’s disease (AD) is a progressive neurodegenerative disorder primarily associated with memory deficits and cognitive decline that can eventually affect behavior, speech, visuospatial orientation and the motor system (1–3). AD is the most common cause of dementia among the elderly, impacting an estimated 32 million individuals globally as of 2023 (4) and its prevalence is increasing in nations with aging populations, thereby imposing significant burdens on their healthcare systems. AD brain pathology is characterized by the accumulation of extracellular amyloid-β (Aβ) plaques (5) and intracellular hyperphosphorylated tau aggregates as neurofibrillary tangles (NFTs) in the gray matter (6). These pathological features can trigger cytotoxic events, neuroinflammation, mitochondrial dysfunction, and contribute to the neuronal stress and degeneration, ultimately resulting in brain atrophy (5, 7). Interestingly, similar pathological hallmarks, can also occur in aged individuals who are not diagnosed with AD (8, 9). The extent of these features can vary greatly between individuals, and they may dominate or be restricted in specific brain regions (8, 9). However, it remains unclear what causes these pathological features and whether they are precursors of neurodegeneration and AD, or simply the products of normal brain aging. Further, while previous studies have proposed that aging is the most profound risk factor for AD (10, 11), the molecular mechanisms underlying the aging-related susceptibility to AD is far from clear (12). Therefore, transcriptome studies of the human brain are crucial for uncovering the molecular mechanisms that differentiate AD from normal aging (NA).

Recent advances in single-cell (sc) or single-nucleus RNA sequencing (sc/snRNA-seq), along with spatial transcriptomics, have revealed remarkable molecular diversity in the cellular landscapes of brains under NA and AD. For instance, the first snRNA-seq study on human AD conducted by Mathys *et al.* (13) demonstrated that in human PFC, the strongest changes associated with AD manifest at the early stage of AD pathological progression and are highly specific to certain cell types. In contrast, genes that are upregulated in the later stages of AD are generally consistent across different brain cell types. Moreover, Chen *et al.* (14) utilized the 10X Visium platform to study the human middle temporal gyrus in three AD cases and three control subjects. By aligning 10X Visium spots (∼55 µm diameter) with adjacent sections stained for Aβ plaques and NFTs, the study observed upregulation of specific AD-related genes and changes in gene co-expression patterns across the bulk of cells near AD pathology, in contrast to those in more distant areas. These studies contributed to a better understanding of how gene regulatory networks drive specific transcriptional changes across various brain cell types, spatial context, and health/physiological conditions, shedding light on possible pathogenic cell subtypes underlying AD and novel cell-cell interactions among neuronal and non-neuronal cells. However, until now, AD-focused sc/snRNA-seq and spatial transcriptomics studies (13–16) lacked the capability to capture spatial information at single-cell resolution. This limitation is particularly significant because even in brains of patients with moderate AD, neurons and non-neuronal cells with normal function may coexist with those affected by AD pathology, and the affected cells may present AD related changes at the molecular level in a cell-type/spatial specific manner. Consequently, case-control studies that rely on sc/snRNA-seq or spatial transcriptomics that do not incorporate single-cell resolution may encounter biases when comparing AD-affected with NA brains. These biases can impede the accurate identification of transcriptomic markers in neurons and glial cells that are influenced by the pathological features of AD in contrast to those resulting from NA brains. Thus, comprehensive understanding of the transcriptional profiles and corresponding spatial information at single-cell resolution is important to unveil the molecular mechanism of AD.

In this study, we are the first to utilize the SpaTial Enhanced Resolution Omics-sequencing (Stereo-seq) (17), a state-of-art spatial transcriptome platform offering large-field-of-view and subcellular resolution to develop a comprehensive, transcriptome-wide, and high definition atlas of the human prefrontal cortex (PFC), a vulnerable brain region in AD (18), across both NA and AD-affected individuals. We mapped the transcriptomic landscape and spatial organization of the neocortex layers and the white matter (WM) in the PFC, revealing detailed pathological alterations in its laminar architecture. Moreover, we identified some novel and unique cellular reactions to both physiological stress from NA and pathological stress from AD, at single-nucleus resolution.

## RESULTS

### The divergence of laminar structure and transcriptional profiles between AD and NA samples

Human postmortem optimal cutting temperature (OCT) embedded samples from the prefrontal cortex (BA10 area) of six AD patients (Braak stages IV-VI, male, aged 69-91) and six male control samples (Braak stages II-III, aged 74-95) were selected for Stereo-seq experiments (see detailed case demographics, clinical and overall neuropathological information in **Table S1**). For each sample, two cryosections (∼10 µm distance between each section) were captured and labeled as No.1 to No.2 from the top to the bottom sections, respectively. After the quality control and area selection on the No.1 hematoxylin and eosin (H&E) sections (**Figure S1**), 10*10 mm area from the No.2 sections were chosen for Stereo-seq profiling. Due to the restricted sampling area on the stereo-seq chip (10*10 mm), the neuropathological findings related to AD across the entire brain may not accurately represent the progression of AD within the specific areas in our spatial transcriptome study. Consequently, to further identify the divergence of transcriptional profiles among different stages of AD, the third sections from each sample were captured and labeled as No.3 (∼50 µm distance between the No.2 and No.3 sections). Based on immunohistochemical (IHC) stains of Aβ plaques (19) on the No.3 sections (**Fig. 1A; Methods**), we classified the AD samples into moderate and severe AD groups. Given the minimal presence of the Aβ plaques in the control samples and the advanced age of these subjects (aged 74-95, older than AD subjects on average), we considered the individuals in control group as NA.

**Fig. 1:**
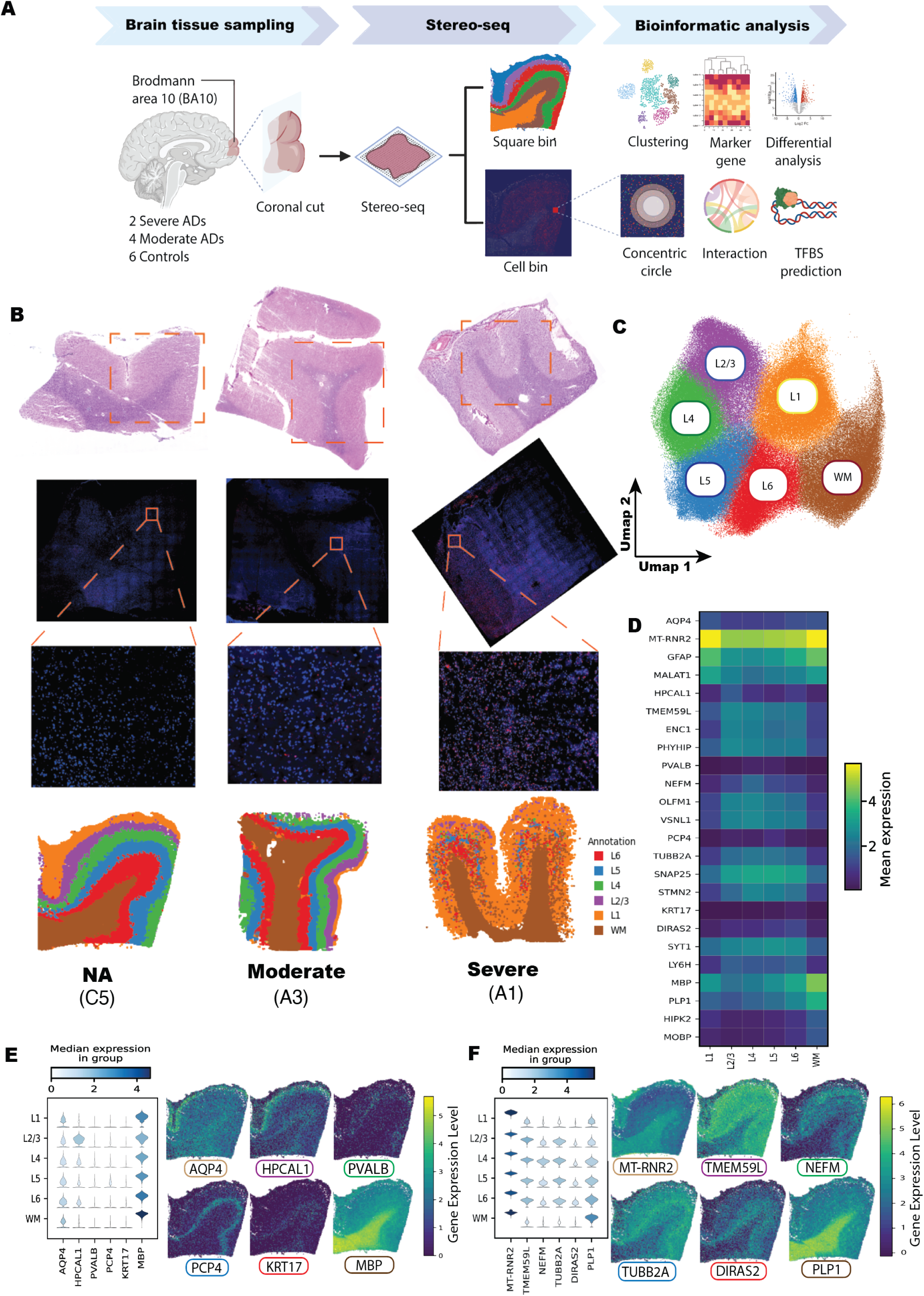
Spatially resolved transcriptomic profiles of the multiple layers in human prefrontal cortex. (A) The analysis pipeline of the study. (B) The H&E staining, immunohistochemistry (IHC) staining for Aβ plaques (Aβ42; red) and nuclei (DAPI; blue), and layer clustering for NA (C5), moderate (A3), and severe (A1) AD groups. (C) UMAP visualization of seven clusters across 12 samples, annotated by cortical layer I to VI and the WM. (D) Heatmap of the marker genes in each cortical layer and the WM. The x-axis represents six cortical layers and the WM. Colors represent the mean expression of the gene in each cortical layer and the WM. (E and F) Violin and scatter plots illustrating the expression levels of previously identified (E) and novel (F) layer marker genes in C5. In the violin plot, the x-axis represents the layer specific markers, and the y-axis represents the cortical layers and the WM. The color represents the median gene expression in the cluster. For the scatter plots, six plots represent the distribution of the gene expression levels of the layer markers, respectively. The color represents the gene expression levels.

Conventionally, the human cerebral cortex is organized into six cellular layers (or laminae) and each layer exhibits unique cellular composition, intra- and interlaminar connectivity, and unique patterns of gene expression (20). A previous spatial transcriptome study by Maynard *et al.* (21) has manually delineated the laminar architecture by aligning gene expression sections with the adjacent H&E sections. They have identified the specific gene markers for each layer in healthy human dorsolateral prefrontal cortex through the 10X Visium platform. However, data-driven unsupervised approaches are crucial for uncovering and understanding the complex structures of the human brain, especially in regions lacking clear histological boundaries. These methods enable the discovery of unknown spatial domains and assist in annotating areas that are difficult to delineate manually, thus playing a key role in brain research. Consequently, in our dataset, to annotate the specific layer based on the layer markers proposed by Maynard *et al* (21) and reveal novel gene markers in each cortical layer, we first converted the raw spatial expression matrix into ∼55µm x ∼55µm pseudo-spots (110 bins x 110 bins/spot, named bin110 resolution), each representing approximately one spot in the 10X Visium platform to facilitate comparable analyses. At the bin110 resolution, a total of 338,410 pseudo-spots were obtained across the 12 samples, comprising 170,465 pseudo-spots from NA samples, 113,324 from moderate AD, and 54,621 from severe AD samples. Each pseudo-spot captured an average of 3,150 counts and 2,225 genes. We then utilized the Harmony algorithm (22) to remove the batch effect of the spatial transcriptional profiles from the 12 samples and employed a newly developed unsupervised spatially constrained-clustering (scc) algorithm (17, 23) across all samples to identify multiple layers in human PFC. Unlike conventional clustering methods for sn/scRNA-seq data (24, 25), scc groups pseudo-spots not only by their gene expression patterns but also by their spatial proximity. This ensures that clusters are formed based on both transcriptional profile similarities and the close spatial arrangement of the pseudo-spots within each cluster (17). In our datasets, the data-driven clustering has successfully identified distinct layers and the WM in all 12 samples based on their unique transcriptional profiles (**Fig. 1B**; **Fig. S1A**). After aligning the gene expression sections with corresponding adjacent H&E sections, we excluded the pseudo-spot without brain tissue or folded regions and plotted all the pseudo-spots on a two-dimensional panel by uniform manifold approximation and projection (26) (UMAP; **Fig. 1B-C; Fig. S1B**). To identify the transcriptional markers of each cortical layer and the WM, we carried out differential gene expression (DGE) analysis by comparing each individual cluster against all the other clusters (**Fig. 1D**; **Methods**). We noticed that specific markers for each layer, *AQP4* in Layer I, *HPCAL1* in Layer II and III, *PVALB* in layer IV, *PCP4* in layer V, *KRT17* in layer VI, and *MBP* in WM (**Fig. 1E**), were consistent with the layer markers proposed by Maynard *et al.* (21), which validated our identified laminar structures of PFC. In addition, we identified multiple novel marker genes for each specific layers across the cortical cortex, including *MT-RNR2* in layer I, *TMEM59L* in layer II and III, *NEFM* in layer IV, *TUBB2A* in Layer V, *DIRAS2* in layer VI, and *PLP1* in WM (**Fig. 1F; Table S2**). Together, our results highlighted the capability of Stereo-seq platform for delineating a high-resolution, fully data-driven spatial transcriptomic atlas of the human brain with transcriptome-wide coverage.

To assess the impact of AD progression on the laminar architecture, we compared the proportion of each layer between samples with different stages of AD. The proportion of each layer is indicative of its thickness relative to the total thickness of the gray matter. Although the layer proportions appeared similar between the moderate AD and NA groups, we noticed a significant reduction in the proportions of layers II-VI in the severe AD samples (**Fig. 2A**). This finding underscores the disruption of laminar architecture and the occurrence of human PFC atrophy in later stages of AD (27). The increased proportion of layer I observed in severe AD samples may be related to extensive neuronal degradation (28) and the high reactivation of astrocytes (Ast) in layers II-VI, a response to the neuronal inflammation (29). Since reactivated Ast in layer II-VI often express genes typically associated with Ast in layer I, such as *GFAP* and *NEAT1* (29) (**Fig. 2B**), this similarity may cause these areas being classified as layer I in our clustering analyses for layers. Furthermore, we conducted the pairwise DGE analysis (**Methods**) among the NA, moderate, and severe AD groups to decipher the AD-related gene expression shifts in different AD stages (**Fig. 2C**). Our results revealed that compared to the NA group, genes down-regulated in the moderate AD group were related to neurotransmission (e.g., *SNAP25*, *GAP43*, *SV2A*, *CLSTN1*) and neuroprotection (e.g., *ENC1*, *APOE*, *OLFM1*), and the upregulated genes were associated with mitochondrial function (e.g., *NDUFA4, LAMP2*), inflammation (e.g., *SPP1*) and myelin sheath structure (e.g., *PLP1*, *CLDN11*). In the severe AD group, besides the genes related to neuronal protection (e.g., *NEAT1*, *MALAT1*), Ast reactivation (e.g., *GFAP*), immune response (e.g., *HLA-B*, *HLA-E*) and blood-brain barrier integrity (e.g., *CLDN5*), we also detected elevated expression of multiple mitochondrial genes compared to the NA group, highlighting mitochondrial dysfunction. Since the mitochondrial energy metabolism is impaired by Aβ toxicity in AD, this up-regulation of mitochondrial genes may represent a compensatory response (30) (**Fig. 2D**). Furthermore, gene ontology (GO) enrichment analysis for the upregulated genes in the moderate and severe AD groups compared with the NA group revealed several AD-related biological processes (**Fig. 2E**). For instance, the Aβ clearance related GO terms, “Positive Regulation Of Macroautophagy”, “Negative Regulation of Amyloid-Beta Formation”, and “Negative Regulation of Amyloid Precursor Protein Catabolic Process”, were enriched in the moderate AD group, and the biological processes of “Positive Regulation of Cell Junction Assembly”, “Presynaptic Membrane Organization”, “Presynaptic Membrane A”, “Synapse Organization”, and “Nervous System Development” were enriched in the severe AD group, suggesting the synaptic dysfunction in the later AD stage. Next, we identified genes that were significantly upregulated in specific cortical layers and the WM within both moderate and severe AD groups, compared to the NA group (**Table S3**). This analysis revealed that unique genes were upregulated across each cortical layer and the WM of the PFC in both moderate and severe AD groups, potentially underlining the basis for different layer vulnerabilities at various stages of AD (31).

**Fig. 2:**
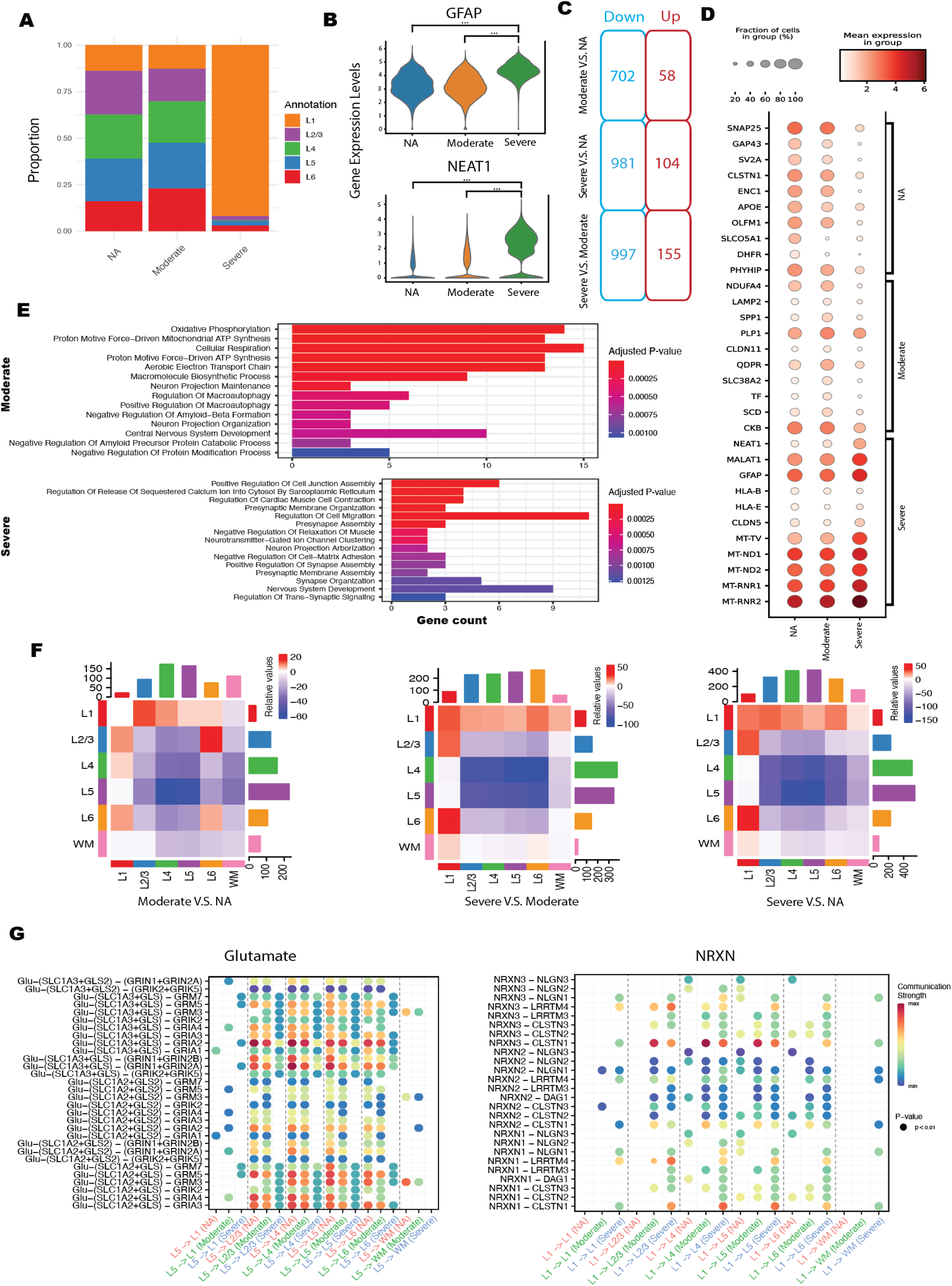
Differences in gene expression patterns between control, moderate AD, and severe AD groups at bin110 resolution. (A) Proportion of the cortical layers in each group. The color represents cortical layers I-VI. (B) The expression level of the *GFAP* and *NEAT1* in NA, moderate, and severe AD groups. (C) The up- and down-regulated genes of the pairwise DGE analysis among all three groups (D) The most significant up-regulated genes in each group after the pairwise DGE analysis among all three groups. Size of the spot indicates the proportion of the cells expressing the genes, and the color represents mean expression level of the genes in each group. (E) GO enrichment analysis based on the up-regulated genes in moderate and severe AD groups compared with the NA group. The length of the bar indicates the gene numbers enriched in the GO term and the color represents the adjusted P-values for enrichment analysis. (F) Pair-wise comparison of the number of outgoing and incoming LR pairs among different cortical layers and the WM across the NA, moderate, and severe AD groups. The color of the squares in the heatmap represents the number of increased (red) or decreased (blue) signaling in the first group compared to the second one. The top color bar represents the total number of changes (increases or decreases) in incoming signaling, and the right color bar corresponds to changes in outgoing signaling, both comparing the first group to the second. (G) The communication strength comparison of the specific LR pairs of Glutamate (from layer V to other layers) and NRXN (from layer I to other layers) signaling pathways across control, moderate and severe AD groups. The x-axis represents the direction of the LR pairs, and the y-axis indicates the specific LR pairs. Dot size represents the adjusted P-value, and the color reflects the communication strength of the LR pairs.

### Layer-layer interaction networks reveal alteration of inter-layer communications in the later stage of AD

The interactions between cortical layers are essential for integrating sensory information, supporting cognitive functions, and facilitating communication within the brain. These interactions involve the vertical transmission of signals, where neurons spanning several layers can integrate inputs and produce outputs (32). In this study, we have constructed the layer-layer communication networks based on the database of interactions among ligands and receptors (33) (**Methods**), aiming to examine the variances in the vertical connections across each cortical layer and the WM between the NA samples and the moderate/severe AD samples. We observed a reduction in the number of interactions across all layers with the progression of AD, decreasing from 3,571 in the NA group to 3,064 in the moderate AD group and 2,244 in the severe AD group (**Fig. S2A**). This pattern indicated the collapse of neural connection networks due to the neurodegeneration in AD, a consequence of Aβ plaques and tau protein aggregation (7). Moreover, we observed a decline in the incoming and outgoing signaling from layers II-VI and the WM, but intensified communications between layer I and the other layers, with the progression of AD (**Fig. S2B**), possibly due to the neurodegeneration in layers II-VI and an increased proportion of layer I in the severe AD group. The layer-layer communication networks in each AD stage showed distinct interaction patterns (**Fig. S2C**). After pair-wise comparisons of the number of interactions among different layers between the NA, moderate, and severe AD groups (**Methods**), we observed that the interactions between the layers IV and V decreased with AD progression, along with the reduction of outgoing interactions from layers IV-V to layers II, III, and VI (**Fig. 2F**). As the layers II-V are crucial in cortical information collection, processing and distributing outputs to subcortical structures (34, 35), the breakdown of the vertical networks leads to the cognitive impairments in AD (36). Compared to the NA group, our investigation further delved into the alterations of the communication strength in each ligand-receptor (LR) gene pair (**Methods**) in moderate and severe AD groups, aiming to reveal the dysregulated inter-layer LR interactions that may be driving the deterioration of cognitive functions. Notable alterations of communication strength related to the Glutamate and neurexins (NRXN) pathways, associated with the neuronal signal transmission (37, 38), were observed between layer IV and V and between layer II and III and V. For instance, the *SLC1A2* and *GLS2—GRIA2* LR pairs, outgoing from layer V to layers II-VI, showed a considerable decline in the moderate AD group and were completely absent in the severe AD group, in contrast to the NA group (**Fig. 2G**). The *SLC1A2*, encoding the excitatory amino acid transporter 2, is a glutamate receptor gene and responsible for clearing the neurotransmitter from the synaptic cleft (39). *GLS2*, encoding glutaminase 2, is essential for maintaining the balance between the production and recycling of glutamate for neurotransmission (40). *GRIA2* encodes subunits of the alpha-amino-3-hydroxy-5-methyl-4-isoxazolepropionic acid (AMPA) receptors, which are integral to fast synaptic transmission in neurons (41). Additionally, we also observed the loss of interactions between *SLC1A2* and *GLS2* ligand genes and other AMPA receptor subunit encoding genes, including *GRIA1, GRIA3*, and *GRIA4*, from layer V to layers II-VI in the severe AD group (**Fig. 2G**). Further, the interaction between *SLC1A2* and *GLS2* ligands and the subunits of metabotropic glutamate receptors (mGluRs), including GRM5 (encoded by *GRM5*) and GRM7 (encoded by *GRM7*) also diminished in the severe AD group. These receptors play crucial roles in the central nervous system by modulating neurotransmitter release and synaptic plasticity, impacting various physiological and pathological processes (42). In addition, from layer I to layers II-V, the expression of *NRXN3*-*CLSTN1*, the ligand-receptor pairs associated with presynaptic differentiation (43), increased in the moderate and severe AD groups, compared to the NA group (**Fig. 2G**). Since presynaptic disruption was previously observed in AD samples (44), the enhancement of these interactions in AD groups may reflect compensatory mechanisms. Together, our results uncovered significant alterations in inter-layer communications within AD samples, particularly in neurotransmission processes. These findings could help elucidate the key mechanisms driving cognitive decline in AD.

### Identification of the location of brain segments under high stress due to NA and AD in human PFC

As the brain ages, neurons become increasingly vulnerable to oxidative stress, inflammation, and protein aggregation (45), contributing to the accumulation of Aβ amyloid into plaques (46) and NFTs formation (47). Additionally, the presence of Aβ plaques and NFTs intensifies neuronal stress, leading to further neurodegeneration (48). Furthermore, Ast and microglia (Mic) surrounding highly stressed neurons become reactivated and play a crucial role in the processes of Aβ clearance and neuroprotection (49). The variations in how neurons and surrounding glial cells respond to stress in both NA and AD remain poorly understood. To investigate this at single-nucleus resolution, we first pinpointed the potential locations of neurons experiencing elevated stress levels in all samples. Considering the heightened vulnerability of excitatory neurons (Ex) to aging (46) and AD (31), coupled with the known positive correlation between cellular apoptosis and elevated mitochondrial gene ratio (number of mitochondrial genes/total detected genes) in cells (48), we presumed that Ex with a high mitochondrial gene ratio may represent Ex under significant stress caused by aging or AD pathological hallmarks, and the positions of these highly stressed Ex mark the potential high stress focal point in human PFC. To precisely identify the location of these highly stressed neurons, we refined our analytic strategy by converting the raw expression matrix into ∼25µm x ∼25µm pseudo-spots (50 bins x 50 bins/spot, named here bin50), which is the default resolution provided by Stereopy (23). Since the mitochondrial genes are exclusively expressed within the mitochondria in the cell cytoplasm (50), pseudo-spots at bin50 resolution, which capture gene expression patterns from both the cell nucleus and cytoplasm, can effectively detect mitochondrial gene expression. This capability aids in accurately identifying pseudo-spots that either fully or partially cover the neurons under high stress. After filtering out the pseudo-spots with low sequencing quality (pseudo-spots with lower than 400 genes detected), we obtained 924,780 pseudo-spots with an average of 834 (and median = 758) genes detected per pseudo-spot. To extract the pseudo-spots partially or fully covering the Ex, we first utilized Harmony (22) to integrate the pseudo-spots from all the samples and then clustered them into multiple distinct subsets based on their transcriptional profiles. After the identification of the marker genes in each cluster, we annotated the clusters as Ast, endothelial cells (End), Ex, inhibitory neurons (Inh), Mic, oligodendrocytes (Oli), and oligodendrocyte progenitor cells (Opc), based on the cell type specific markers (e.g., *AQP4* in Ast, *CLDN5* in End, *SNAP25* in Ex, *GAD1* in Inh, *CD74* in Mic, *MBP* in Oli, *LHFPL3* in Opc) proposed by a previous snRNA-seq study on human PFC (13) (**Fig. 3A-B; Table S4**). To check the accuracy of our annotation results, we aligned the annotated bin50 pseudo-spots with corresponding adjacent H&E sections. Ex were predominantly situated in the neocortex, while a high concentration of Oli was observed in the WM (**Fig. 1B**, **Fig. 3C**, and **Fig. S1A**), which strongly support that although the pseudo-spots were not at single-nucleus resolution, our results still accurately delineated the proximate positions of the Ex. In addition, we noticed that in moderate and severe AD groups, and especially in the severe AD group, a large proportion of Ex and Oli were filtered out due to the low gene number detection (**Fig. 3C** and **Fig. S3A**), reflecting the Ex degradation (28) and changes in the WM caused by Wallerian-like degeneration (51) in AD.

**Fig. 3:**
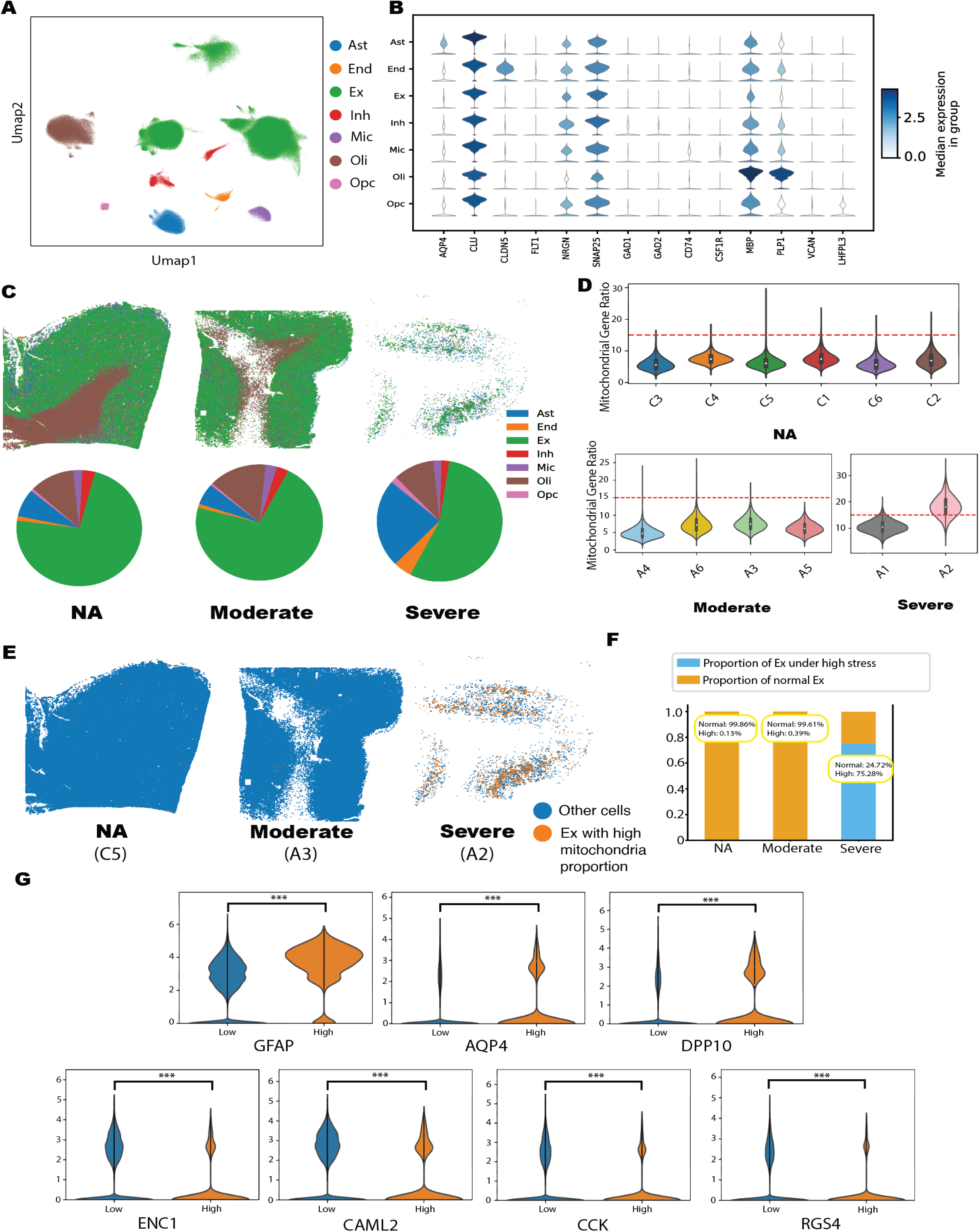
Identity of the location of the dysfunctional neurons in the three groups at bin50 resolution. (A) UMAP visualization of pseudo-spot clusters. (B) Expression levels of the gene markers in each cluster. The color of the violin plot reflects the median expression of the gene in each cluster. Based on the gene markers, we annotated the clusters as astrocytes (Ast), endothelial cells (End), excitatory neurons (Ex), inhibitory neurons (Inh), microglia (Mic), oligodendrocytes (Oli), and oligodendrocyte progenitor cells (Opc). (C) Distribution of different cell clusters on the captured area of Stereo-seq platform (C5 in NA, A3 in moderate, and A2 in severe AD group), and the proportions of cell clusters in all samples from the NA, moderate, and severe AD groups, respectively. (D) The violin plot of the mitochondrial gene ratio of Ex in each sample. The x-axis represents 12 samples and the y-axis indicates the mitochondrial gene ratio. The dashed line represents the threshold of the mitochondrial gene ratio (15%) to classify the Ex into highly stressed or normal ones. (E) Distribution of the highly stressed Ex in NA, moderate, and severe AD groups. The yellow dots represent the pseudo-spots covering the highly stressed Ex, and the blue dots are pseudo-spots containing the normal Ex and other types of the cells (Ast, End, Inh, Mic, Oli, and Opc). (F) Proportion of the highly stressed Ex pseudo-spots among all Ex pseudo-spots in NA, moderate, and severe AD groups. (G) The significant up- and down-regulated genes of the highly stressed Ex pseudo-spots, compared to normal Ex pseudo-spots.

Due to the lack of an established mitochondrial gene ratio threshold for distinguishing stressed Ex from healthy Ex in spatial transcriptome data, we set a threshold at 15% based on the distribution of mitochondrial gene ratio across the pseudo-spots covering the Ex from all the samples (**Fig. 3D**), that is, pseudo-spots annotated as Ex that exhibit ≥15% mitochondrial gene ratio were classified as containing neurons under high stress. In the NA group, we noticed that only 0.13% of the Ex pseudo-spots contain Ex under high stress, indicating minimal high-stress focal points in NA. In contrast, the proportion of Ex under high stress was slightly increased in the moderate AD group and was dramatically higher in the severe AD group (**Fig. 3E-F**), highlighting the prevalence of Ex with high stress in the later stage of AD. We further checked the DEGs between the pseudo-spots covering highly stressed neurons and those containing the normal ones. Besides the mitochondrial genes (e.g., *MT-RNR2*, *MT-ND4*, and *MT-CYB)*, multiple AD-related genes, including *GFAP* (52), *AQP4* (53), and *DPP10* (54), were also upregulated in the pseudo-spots covering highly stressed Ex, compared to those containing normal Ex (**Fig. 3G**). In the pseudo-spots covering highly stressed neurons, the high expression of Ast markers, *GFAP* and *AQP4*, indicated that these areas also include the transcriptional profiles of adjacent Ast. This observation reflects that when neurons are subjected to high stress, the nearby astrocytes become reactivated and migrate towards these neurons to support their survival (55). Meanwhile, consistent with previous transcriptional studies in AD (56–59), *ENC1*, *CALM2*, *CCK, RGS4* were downregulated in the pseudo-spots covering highly stressed Ex (**Fig. 3G**). Together, these findings support our hypothesis that pseudo-spots containing Ex with elevated mitochondrial gene ratio represent brain areas under high stress affected by NA or AD pathology hallmarks.

### Transcriptional divergence at single-nucleus resolution between brain cells impacted and not impacted by NA and AD pathology hallmarks

To further delineate the transcriptional landscape at single-nucleus resolution, we applied a deep learning model (23) for the nucleus segmentation to delineate the cell nuclei based on the nuclei staining image (**Methods**). After filtering out the low-quality nuclei (i.e., nuclei with < 150 detected genes), we analyzed 398,741 nuclei, with an average of 524 (and median=398) genes detected per nucleus. We performed clustering analysis to identify the nuclei subgroups (**Fig. 4A**) and conducted pairwise DGE analyses among the subgroups to identify the specific marker genes for each nucleus cluster. Based on the marker genes in each cluster, we successfully annotated nuclei as the Ast (*AQP4, GLUL*), End (*CLDN5, IFITM2*), Ex (*NRGN, SYT1*), Inh (*GAD1, GAD2*), Mic (*CD74, CD14*), Oli (*MBP, PLP1*), and Opc (*VCAN*) (13) (**Fig. 4B** and **Fig. S4A**). The high distribution of the Oli in the WM and Ex in gray matters substantiated the accuracy of our annotation (**Fig. 4C and 1B**).

**Fig. 4:**
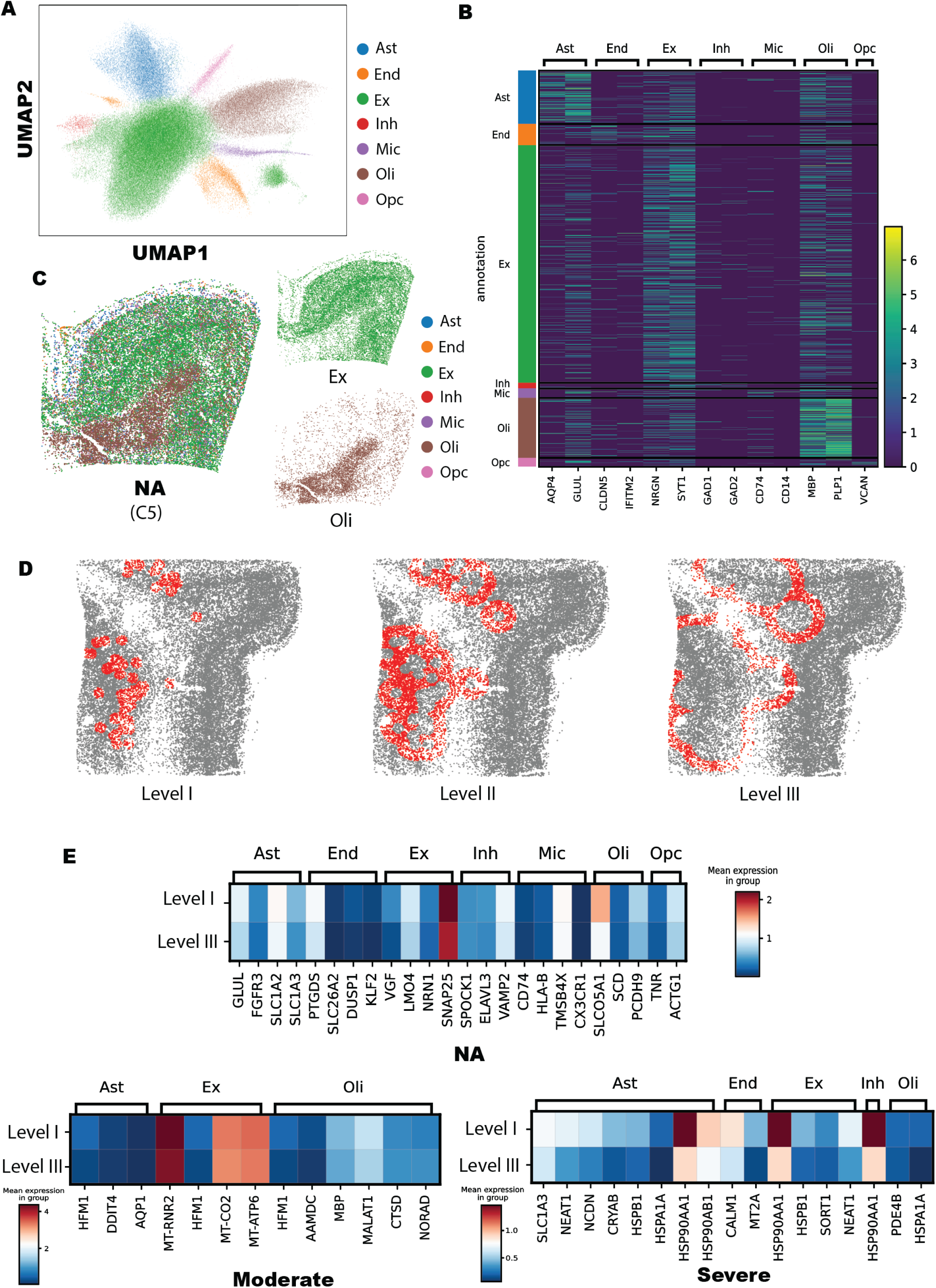
Concentric analysis at the single-nucleus resolution. (A) UMAP visualization of clusters of nuclei. (B) Heatmap of the gene marker expression in each cluster of nuclei. The color represents the expression levels of the gene in each cluster of nuclei. (C) The nuclei type distribution on C5 (NA) sample. (D) Distribution of nuclei in A3 (moderate) across levels I, II, and III in the concentric circle analysis. The red dots represent the selected nuclei at each level, and gray dots indicate other nuclei. (E) The nucleus-type specific DGE analysis between the nuclei in level I and III across control, moderate, and severe AD groups. The color represents the mean expression of the genes in each level.

Compared to the healthy neurons, those experiencing high stress due to NA and AD pathology hallmarks exhibit distinct transcriptional profiles as part of their stress responses (60, 61). Additionally, the mechanisms of these stress responses can differ between neurons affected by NA and those influenced by AD pathology (62). Furthermore, glial cells located near the areas under high stress are more actively involved in neuroprotective actions to aid neuronal survival, in contrast to those situated farther away (55, 63). Thus, to enhance our understanding of the cellular responses to the stress caused by NA and AD pathology, we extracted the nuclei within approximately 1,250 µm from the location of high stress focal point (identified at bin50 resolution) across all samples, totaling 86,626 nuclei, and performed concentric circle analysis (**Methods**). Briefly, the extracted nuclei were classified into three groups (levels I to III) according to their proximity to the high-stress focal point (**Fig. 4D**). Considering that neuron somas range from approximately 4∼100 µm in size, to fully include a whole stressed neuron and the nearby glial and endothelial nuclei, we categorized nuclei located within 250 µm to the stressed neurons as level I, 250-750 µm as level II, and 750-1,500 µm as level III. Given the close proximity and highly similar transcriptional profiles between nuclei in levels I and II, we focused the nuclei type specific DGE analysis between nuclei in levels I and III across NA, moderate and severe AD groups, respectively, in order to distinguish gene expression patterns between nuclei that are close to and those that are more distant from stressed neurons at different stages of AD. Our findings indicat that as with progression of AD, there was notable nucleus type specific differentiation in gene expression patterns between nuclei of level I and level III (**Fig. 4E**; **Table S5**). For instance, in the NA group, Ex at level I significantly increased the expression of neuroprotective genes, including *VGF* (64), *LMO4* (65), and *NRN1* (66), when compared to level III. This highlights Ex’s mechanisms in mitigating stress from NA, thereby enhancing neuron survival. However, this pattern of upregulation for *VGF*, *LMO4*, and *NRN1* in Ex at level I, in contrast to level III, was not present in the moderate and severe AD groups. In the severe group, compared to level III, Ex at level I notably enhanced the expression of genes for heat-shock proteins (*HSP90AA1* and *HSPB1*) as a reaction to neuroinflammation and neurotoxic substances (67); additionally, there was a significant increase in the expression of genes associated with the clearance of Aβ peptides, such as *SORT1* (68). The long non-coding RNA (lncRNA) *NEAT1,* which contributes to neuron damage by downregulating microRNA-27a-3p (69), was also significantly upregulated in level I Ex in the severe AD group, in contrast to Ex in level III. In the NA group’s Ast, when compared to those in level III, there was a notable increase in the expression of *SLC1A3* in the level I Ast. This gene is known for its role in protecting neurons against glutamate-induced excitotoxicity by clearing excess glutamate from the synaptic gap (70). Like Ex, Ast in the severe group at level I, in comparison to level III within the same group, also showed a marked increase in the expression of heat-shock protein genes (e.g., *CRYAB, HSPB1, HSPA1A, HSP90AA1, HSP90AB1*). Furthermore, unlike the nuclei type-specific upregulated genes observed in level I compared to level III in the moderate group, there is a notable upregulation of heat-shock protein-coding genes in Ast, Ex, Inh, and Oli in level I compared to level III in the severe group. This observation aligns with the findings proposed by Mathys *et al.* (13), which suggest that AD-associated changes manifest early in the pathological progression and are highly cell-type specific, while genes upregulated in later stages are common across cell types and predominantly involved in the global stress response. Together, our results demonstrated the intricate dynamics and differences of cellular responses to NA and AD, revealing distinct cell-specific gene expression alterations linked to the spatial position relative to stressed neurons.

### Cell-cell communication networks reveal the reduction of neuron-protective LR interactions in cells near stress-affected neurons in AD

The collaborative mechanisms between neurons and glial cells play a vital role in inhibiting the accumulation of Aβ and the aggregation of tau proteins (71). While prior snRNA-seq studies (72, 73) have uncovered distinct intercellular interactions in AD cases and NA controls, the spatial information, particularly the proximity of glial cells and End to the neurons under high stress in AD and NA, has been largely ignored. As a result, to assess whether neurons under stress and their surrounding glial cells and End employ similar or distinct mechanisms in response to the stress caused by NA and AD pathology, we conducted cell-cell communication analysis (33) across nuclei within levels I and III for all three groups (NA, moderate, and severe AD groups), (**Fig. 5A-B**). We found that the level I nuclei showed higher number and intensity of LR pairs than the level III nuclei in the NA group (**Fig. 5A**), suggesting that intercellular interactions among the neurons, glial cells, and End were enhanced in cells proximal to stressed neurons during NA, potentially as a protective mechanism against the degradation of stressed neurons (**Fig. 5B**). However, as AD progressed, the communication networks among level I cells became compromised, indicating that the intercellular interactions close to stressed neurons are disrupted in moderate and severe AD groups (73) (**Fig. 5A-B**). We further identified some specific LR pairs exclusively activated in the level I nuclei of the NA group, including some that are known to play crucial roles for neuroprotection and Aβ clearance (**Fig. 5C**). For instance, the PSAP (encoded by *PSAP*) released by Ex, End, Mic, Oli, and Opc acts on the GPR37L1 (encoded by *GPR37L1*) on Ast to activate the motility of Ast and release of diffusible neuroprotective factors to shield the neurons affected by neurotoxic damage(74). The Amyloid Precursor Protein (encoded by *APP*) released by Ex and Inh, interacts with the CD74 (encoded by *CD74*) on Mic to inhibit Aβ production (75). These interactions were impaired or diminished in the nuclei proximal to the stressed neurons (level I) in moderate and severe AD groups (**Fig. 5C-E**). Together, our results suggest that, compared to NA, highly stressed neurons and surrounding glial cells in AD samples lose their ability for Aβ clearance and neuroprotection. This loss can contribute to the formation of Aβ plaques and exacerbate neuronal degradation, potentially explaining the high levels of Aβ deposition and brain atrophy observed in AD.

**Fig. 5:**
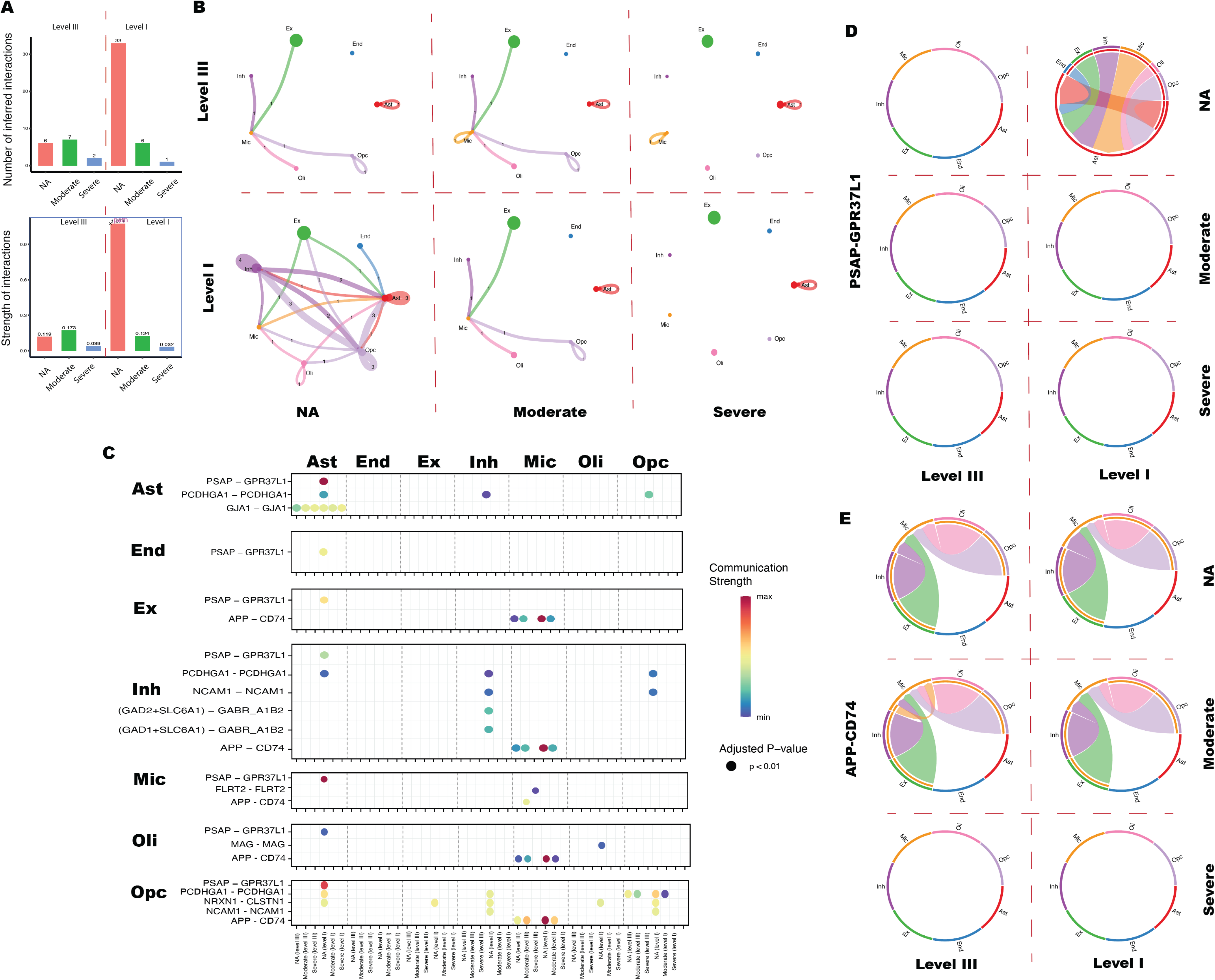
Differences in the cell-cell communication between the nuclei in level I and III. (A) The total number (top) and the strength (bottom) of the LR pairs in level I and III groups across control, moderate, and severe AD groups, respectively. (B) Networks of cell-cell interactions show the number of LR pairs (edges) between nuclei clusters (nodes) within level III and I across NA, moderate, and severe AD groups, respectively. The colors of the dots and edges represent the specific types of nuclei and the outgoing signaling emanating from them. The number on the edge indicates the number of the outgoing signals. The size of the spot are proportional to the number of nucleus within each level of specific AD group. (C) The communication strength of the specific LR pairs of the nuclei in level I and III in the NA, moderate, and severe AD groups, respectively. Nuclei type in each row indicates the nuclei expressing the ligands and the column indicates the one expressing the receptors. The x-axis represents the level I and III across NA, moderate, and severe AD groups in each nucleus type. The color of the dots reflects the communication strength, and the size of the spot represents the adjusted P-value. (D-E) The chord diagram of the PSAP-GPR37L1 and APP-CD74 LR pairs of the nuclei in level I and III in the NA, moderate, and severe AD groups, respectively. Each segment of the circle in different color represents different types of nuclei. The chords connecting the segments represent interactions between two types of nuclei, and the thickness of each chord reflecting the strength of the interactions.

### Co-expression networks at single-cell resolution uncover key gene markers and potential regulatory transcription factors in AD progression

Although our results uncovered nucleus-type specific changes associated with brain areas under high stress caused by NA and AD pathology, the complex relationships and patterns among co-expression gene modules, as well as the key regulators involved in the neuron protection and the clearance of AD pathological hallmarks, remain unclear. Using hdWGCNA (76), we constructed the nucleus type specific weighted gene co-expression networks on the 2,000 most variable genes for each nucleus type from levels I-III across all samples. We successfully identified two co-expression gene modules in Ex and Inh, respectively (**Fig. 6A, Fig. S4B-C**), but not in the other nucleus types due to the low sequencing depth and limited nuclei numbers. We noticed that in these modules, several hub genes have been reported to be related to AD. For instance, in the Ex1 module, the hub gene *UCHL1,* encoding the ubiquitin C-terminal hydrolase L1 protein, is a major neuronal enzyme involved in the elimination of misfolded proteins (77). The decreasing expression of this gene is responsible for Aβ42 accumulation (77). Additionally, within the Ex1 module, we identified two hub genes, *ENC1* and *RTN1*, which are key genes implicated in the transition from asymptomatic to symptomatic AD (78). In the Inh1 module, the hub gene *VAMP2* is known to be related to neurodegenerative disease where a reduction of *VAMP2* expression was associated with cognitive decline (79), and another hub gene, *PRNP*, which encodes prion protein, is involved in neuroprotection to excitotoxicity (80) and has inhibitory effects on BACE1 to decrease A□ production (81). We also noticed that one hub gene in the Inh2 module, *HSPA8* (encoding heat-shock protein family A member 8), can directly disassemble RHIM-amyloids to inhibit necroptosis signaling in cells and in mice (82). To enhance our understanding of the biological functions of all four modules (Ex1, Ex2, Inh1, Inh2), we conducted GO enrichment analysis on the top 50 hub genes based on connectivity within each module. Our results indicate that all four modules are primarily associated with neuroprotective processes and inhibition of the formation of AD pathological hallmarks. For instance, the Ex1 module was significantly enriched in processes such as “Negative Regulation of Amyloid-Beta Formation”, “Negative Regulation of Protein Phosphorylation”, and “Negative Regulation of Peptidyl-Threonine Phosphorylation” (**Fig. 6B**), suggesting its critical role in mitigating the formation of AD pathological hallmarks. Besides the “Negative Regulation of Amyloid-Beta Formation” GO term, the Ex2 module also supported processes like “Negative Regulation of Intrinsic Apoptotic Signaling Pathway” and “Negative Regulation of Neuron Death” (**Fig. S4D**), which may help in preventing the stressed neurons from apoptosis. The Inh1 module was mainly enriched for processes such as “Positive Regulation of Synaptic Transmission”, “Negative Regulation of Cytokine Production”, “Response to Amyloid-Beta”, and “Regulation of DNA Repair” (**Fig. 6C**), while the Inh2 module was also enriched for terms related to Aβ clearance (**Fig. 6D**). These results underscore the involvement of these modules in neuroprotective responses against aging and AD-induced stress. Additionally, we performed module-trait correlation analyses to assess the correlations of each module with AD progression (NA, moderate AD, and severe AD groups) and with spatial proximity to stressed brain areas (levels I-III groups). While the Ex2 module showed low correlation with AD progression (r=0.0261, adjusted P-value<0.01) (**Table S6**) or spatial proximity to the stressed neurons (r=-0.0423, adjusted P-value=0.0675) (**Table S6**), the Ex1 module exhibited negative correlations both with AD progression levels (*r*=-0.2241, adjusted P-value<0.01) and spatial proximity to stressed neurons (r=-0.0921, adjusted P-value<0.01) (**Table S6**). Moreover, the Inh1 and Inh2 modules both showed significant negative correlations with AD progression (Inh1: r=-0.2193, adjusted P-value<0.01; Inh2: r=-0.2577, adjusted P-value<0.01) (**Table S6**). This implied a diminished capacity in Ex to inhibit the formation of Aβ plaques and tau hyperphosphorylation when subjected to heightened stress in AD samples. Intriguingly, the Inh1 module also demonstrated a positive correlation with spatial proximity to high stressed neurons (r=0.1196, adjusted P-value<0.01; **Table S6**), suggesting Inh close to the stressed brain areas may have an enhanced neuroprotective capability to support the Ex survival and maintain the Ex/Inh balance in brain (83).

**Fig. 6:**
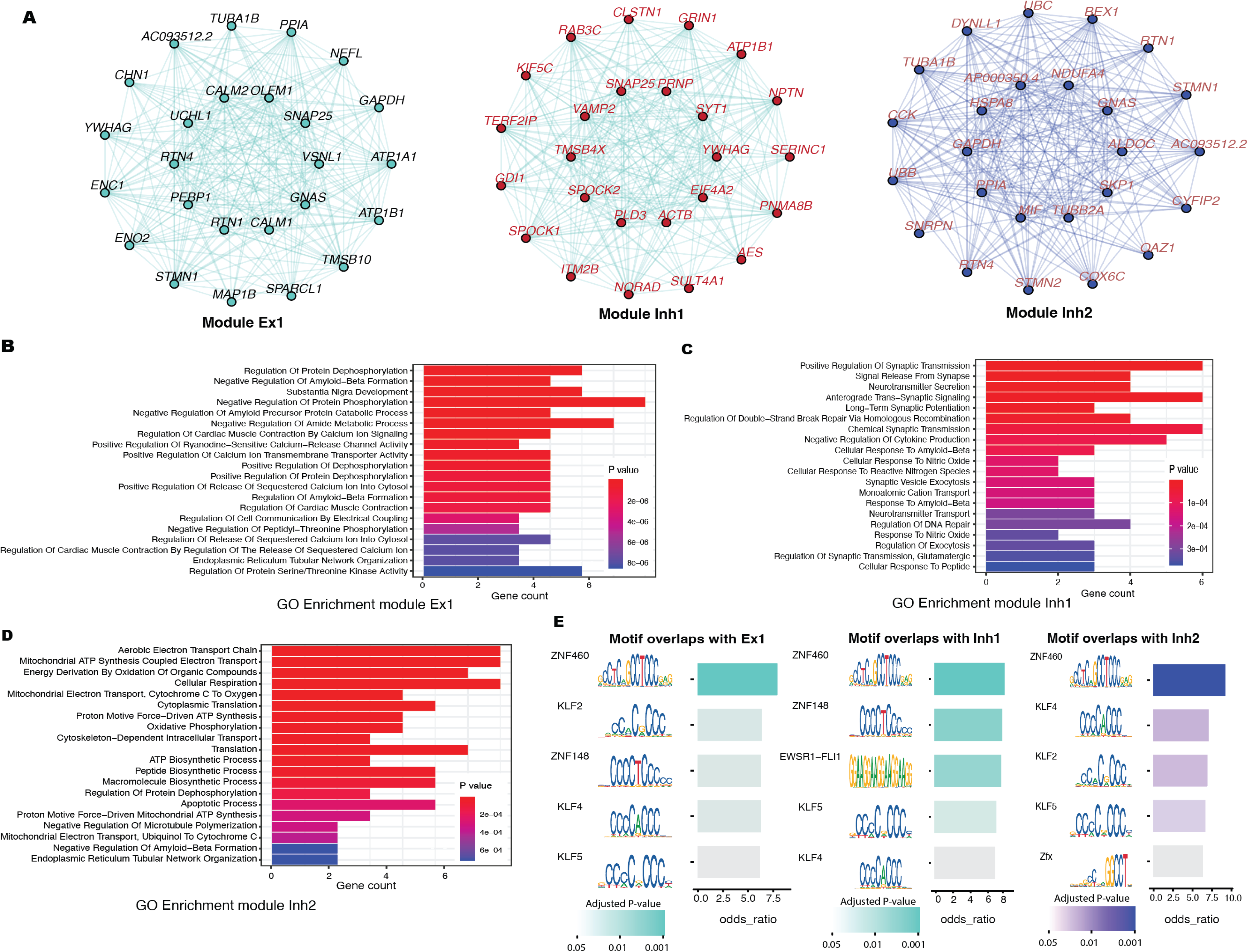
Gene co-expression networks in excitatory and inhibitory neurons. (A) The gene co-expression modules in Ex (Ex1) and Inh (Inh1 and Inh2). The nodes represent the hub genes, while the edge connecting two nodes indicates the co-expression of those genes. (B-D) GO Enrichment analysis on the top 50 hub genes in the Ex1, Inh1, and Inh2 modules. The length of the bar indicates the gene numbers enriched in the GO term and the color represents the adjusted P-values for enrichment analysis. (E) Enrichment analysis to identify the Motif overlaps with Ex and Inh modules. The color indicates the adjusted P-value.

Given that the three modules (Ex1, Inh1, and Inh2) were negatively correlated with AD progression, the transcription factors (TFs) regulating these modules could serve as potential therapeutic targets to prevent the formation of AD pathological hallmarks and neuronal degradation in AD (84). To this end, we utilized the MotifScan package (85) to find the TFs significantly enriched within each module (**Methods**). We identified that KLF2 was significantly enriched in module Ex1 and Inh2 (**Fig. 6E**). Fang *et al.* (86) have proposed that the upregulation of Kruppel-like factor 2 (encoded by *KLF2*) attenuates oxidative stress triggered by Aβ, improves mitochondria function, and reduce the rate of apoptosis. In addition to the neuroprotective TF, we noted that two TFs, KLF4 and KLF5 were significantly enriched in all the three modules (**Fig. 6E**). These two TFs are recognized for accelerating the progression of AD. Specifically, KLF4 was identified as a key mediator in promoting Aβ-induced neuroinflammation by exacerbating the release of pro-inflammatory factors (87), and KLF5 can accelerate APP amyloidogenic metabolism and promote Aβ synthesis through binding to the BACE1 promoter (88). Interestingly, one TF, ZNF460 showed the highest level of enrichment across Ex1, Inh1, and Inh2 gene modules (**Fig. 6E**), suggesting its potential role as a pivotal regulator in neuronal stress responses to NA or AD, as well as in neuron degradation processes.

## DISCUSSION

In this study, the Stereo-seq platform (17), a high-resolution spatial transcriptome technology, was employed for the first time to develop the most comprehensive, data-driven, transcriptome-wide molecular atlas of the adult human PFC ever achieved. Additionally, with the unparalleled resolution and the largest sample size in spatial transcriptomics for human PFC to date, including six AD cases and six NA controls of the same sex and approximately matched by age with slightly older controls, our study uncovered significant molecular alterations in AD samples relative to NA controls at both cortical layer and single-nucleus levels. By analyzing high-definition landscapes of the human PFC in both AD cases and NA controls, we have identified the transcriptional and structural alterations across six cortical layers and the WM in PFC associated with AD. Further, we have unveiled unique stress response mechanisms among neurons and adjacent non-neuronal cells, both during NA and throughout various stages of AD.

Here, we highlight some of the novel findings. First, at bin110 resolution, our results demonstrated the change in laminar architecture with the progression of AD. While the proportion of the cortical layers remained the same in the moderate AD group compared with the NA group, layers II-VI were diminished in the late stage of AD. This reflects the atrophy of the PFC attributed to neuronal degradation (89) and the reactivation of the Ast in response to toxic substance in the brain displahing AD pathology hallmarks (90). Further, disruptions in the neocortex’s laminar architecture led to impaired interactions between layers in AD samples compared to NA controls. These interactions included the Glutamate and NRXN signal pathways which play critical roles in neurotransmission (37, 38), indicating the AD pathology hallmarks break down the connection of neurons in different layers and lead to damage in cognitive function.

Since the cellular responses to the stress caused by oxidative damage, inflammation, and protein aggregation are the key factors leading to Aβ plaque deposition (91) and NFT formation (47). We uniquely identified distinct stress response mechanisms at the single-nucleus level in neurons and adjacent glial cells, comparing the responses between NA and AD. In the NA group, our analysis revealed amplified *CD74-APP* LR pairs between stressed Ex and adjacent Mic which may enhance Aβ clearance (75). The motility of the nearby Ast was also activated through *PSAP-GPR37L1* pairs, prompting their migration towards stressed neurons to offer protection (74). However, these neuron protective LR pairs were found to be compromised in moderate and severe AD groups when neurons are under stress. In summary, our results suggest that the impaired capability of Aβ clearance and neuroprotection are key factors in the deposition of Aβ plaques in AD brains, aligning with the findings proposed by Wildsmith *et al.* (92). Moreover, further accumulation of Aβ may lead to neuronal degradation and ultimately contribute to the progression of AD.

To further support our findings that a decrease in Aβ clearance contributes to the progression from NA to AD, we have conducted additional gene co-expression analysis. The nucleus-type specific gene co-expression networks revealed that in the Ex and Inh, the three gene modules, Ex1, Inh1, and Inh2, related to the Aβ plaque and NFT clearance progressively decreased during AD progression. Our findings indicate that while stressed neurons contribute to the accumulation of Aβ into plaques (46) and the formation of NFTs (47), in the context of the NA group, both stressed neurons and adjacent brain cells participate in the clearance of Aβ plaques and the dephosphorylation of proteins, which prevents the accumulation of Aβ plaques and tau aggregation, respectively. Conversely, in AD brains, this protective mechanism is compromised. Stressed neurons and surrounding cells lose their capacity to clear Aβ and maintain protein dephosphorylation, leading to the formation of AD pathology hallmarks and acceleration of AD progression. These observations align with previous research indicating that impaired Aβ clearance is a hallmark of AD samples (93, 94) and a decrease in tau phosphatase activity in AD brains contributes to an imbalance in the protein phosphorylation/dephosphorylation system, culminating in NFT formation (95). Interestingly, one transcript factor, ZNF460, is a regulator with the highest enrichment in all three modules (Ex1, Inh1, and Inh2), and thus may be an important therapeutic target to promote neuron survival and AD pathological hallmarks clearance. Although the biological mechanisms linking ZNF460 to AD has yet to be established, a study by Liu *et al.* (96) demonstrated that ZNF460 interacts with the apolipoprotein C1 (APOC1) promoter and enhances APOC1 transcription. This process is believed to contribute to the progression of gastric cancer (96). Notably, APOC1, like apolipoprotein E (APOE), is involved in lipid metabolism (97), and its H2 allele has been identified as a genetic risk factor for AD (98). Future studies are needed to explore whether a similar interaction between ZNF460 and APOC1 occurs in the brain, and to assess its potential as a therapeutic target for AD. Overall, our study offers insights into the potential mechanisms underlying the progression of AD from NA to its late stages, and identified one key TF, ZNF460, that may regulate these mechanisms.

Despite the distinct advances and novel findings that this study contributes to the field, our study may also have some limitations that future studies with more advanced technologies and clinical samples may address. First, although Stereo-seq technology offers us the highest resolution among the current spatial transcriptome platforms, the average number of genes captured per cell (∼500 median genes per cell) is significantly lower than that achieved by traditional snRNA-seq methods. While we successfully identified neurons, glial cells, and End at single-nucleus resolution, the reduced sequencing depth may potentially introduce bias in detecting significant alterations in certain transcriptomic molecules related to AD. Enhancements in mRNA capture efficiency could improve the sensitivity and accuracy of spatially resolved cellular taxonomy within the brain. Additionally, our study exclusively involved male samples, which might lead to bias when considering the impact of AD on females. Despite these challenges, our findings offer crucial and valuable insights into the transcriptional landscape of AD on the human PFC at various resolutions, and they provide a detailed and systematic understanding of alterations in neocortical laminar architecture and specific stress responses at various stages of AD, compared to the NA.

## MATERIALS AND METHODS

### Study Subjects

A total of 12 postmortem samples from the prefrontal cortex (Brodmann area 10) were collected at the University of Kansas (KU) AD Research Center (ADRC). These included samples from six AD cases, three samples of primary age-related tauopathy (PART), and three samples exhibiting low AD burden. All samples were from Non-Hispanic White males age 70-95 years. Given that cognitive decline in individuals with PART and low AD burden may not be significant or even detectable (8), we classified these six samples as controls. Informed consent was approved by the KU Institutional Review Board (IRB) and obtained for all human participants. For details on human samples used in this study, please see Supplementary Table 1.

### Tissue Preparation, Cryosection, Stereo-seq Library Preparation, and Sequencing

Stereo-seq is a state-of-art spatial transcriptome platform that captures mRNA from tissue sections using stereo chips. This technology achieves nanoscale resolution with a spot diameter of 220nm, enabling the most detailed delineation of the transcriptome landscape currently available. Further, distinct from other spatial transcriptome platforms, the Stereo-seq chip has the capability to convert a few hundred spots into a pseudo-spot by combining the transcriptional information from the selected DNA nanoball (DNB) patterned array (17), providing us the opportunity to depict transcriptional profiles at multiple resolutions.

To construct a comprehensive spatial transcriptome atlas of human PFC, the BA10 area from 12 postmortem human brains was harvested and immediately flash frozen, embedded with Tissue-Tek OCT medium (Cat # 4583, SAKURA FINETECK USA Inc.) in liquid nitrogen, and then stored at −80°C until ready for the Stereo-seq pipeline. Cryosections were cut at a thickness of 10 µm and mounted on Stereo-seq permeabilization chips (Cat # 210CP118, STOmics America Ltd) or transcriptomics chips (Cat # 210CT114, STOmics America Ltd). Tissue fixation and the following spatial transcriptomics procedures were performed according to the vendor’s manual and previous publications (17, 99). In brief, the tissue section on the Stereo-seq chip (10 mm x 10 mm) was incubated at 37°C for 5 mins, and subsequently fixed in pre-cooled methanol (Cat # 34860, Sigma) at −20°C for 30 mins. Once the fixation was completed, the chip was placed under a ventilation hood to allow residual methanol to air dry. The tissue section on the chip was then stained with nucleic acid reagent (Cat # Q10212, 0.5% v/v, Invitrogen) for 5 mins and subsequently washed with 0.1X SSC buffer (Cat # AM9770, 0.05 U/mL RNase inhibitor, Ambion). The nuclei images were captured using a Zeiss Axio Scan Z1 microscope (at EGFP wavelength). Subsequently, the tissue section was incubated in the permeabilization buffer (Cat # 111KP118, STOmics America Ltd) for 12 mins at 37°C. Stereo-seq transcriptomics chip-captured RNAs from the permeated tissue were then reverse transcribed for 3 hours at 42°C. Next, the tissue was removed and the cDNAs were released from the chip using the transcriptomics reagent kit (Cat # 111KT114, STOmics America Ltd). After the cDNA obtained was size-selected, amplified, and purified, the concentration was quantified by Qubit dsDNA HS assay kit (Cat # Q32854, Invitrogen). Next, 20 ng of cDNA from each sample were used for library construction using the library preparation kit (Cat # 111KL114, STOmics America Ltd) and subsequently for DNB generation. Finally, the DNBs were sequenced on the DNBSEQTM T7 sequencing platform (Complete Genomics, San Jose, USA) with 50 bp read1 and 100bp read2 (Cat # 100008555, Complete Genomics).

### Tissue Preparation, Cryosection, and Immunohistochemistry

Cryosections were cut at a thickness of 10 µm and mounted onto slides. The sections were fixed and permeabilized with pre-cooled acetone at −20°C for 10 mins, and then rinsed three times with 1X PBS. Sections were placed under a vented hood for air drying prior to AT8 and Aβ42 staining. Phospho-Tau (Ser202, Thr205) Monoclonal Antibody (AT8) (Cat # MN1020, Invitrogen), beta Amyloid (1–42) Polyclonal Antibody (Cat # 44-344, Invitrogen), and Tyramide SuperBoost^TM^ Kit (Cat # B40915, Cat # B40962, and Cat # B40953, Invitrogen) were used for immunohistochemical staining according to the vendor’s instructions (Invitrogen).

### Stereo-seq data processing

The fastq files from Stereo-seq were processed following the standard pipeline (https://github.com/STOmics/SAW). Initially, the first reads containing coordinate identity (CID) sequences underwent alignment to the designed coordinates of the Stereo-seq chip, based on the results from the first round of sequencing. This step allowed for a maximum of one base mismatch during the alignment process to account for potential sequencing errors. After alignment, reads exhibiting molecular identifiers (MIDs) possessing more than two bases with a quality score below 10 were excluded to ensure data integrity. The associated CID and MID for each qualified read were then incorporated into the read header. Retained reads were aligned to the reference genome using STAR (100) and only reads achieving a mapping quality score above 10 were considered for gene annotation, ensuring the accuracy of gene expression profiling. The unique molecular identifiers (UMIs) sharing identical CIDs were combined into a single UMI. This step allowed for a single mismatch, facilitating the correction of sequencing and PCR errors. Finally, the CID-containing expression profile matrix was constructed.

### Bioinformatics analysis of Stereo-seq dataset

#### Stereo-seq data integration and dimension reduction

At the bin110, bin50, and single-nuclei resolution, all the pseudo-spots/nuclei across 12 samples were merged and the 2,000 genes with the highest dispersion (variance/mean) were selected for principal component analysis (PCA). To remove potential batch effects, we have applied the Harmony algorithm (22) to transform the top 50 PCs of the pseudo-spot/nuclei and project all pseudo-spot/nuclei into a shared embedding panel. UMAP (26) was used to project the transformed PCs into a two-dimensional panel.

#### Clustering analysis

To identify the laminar architecture of human PFC at the bin110 resolution, the spatial constrained clustering algorithm was employed using the ‘tl.spatial_neighbors’ and the ‘tl.leiden’ functions in Stereopy package (v1.0.0) (23) with the default parameter. At the bin50 and single-nucleus resolution, ‘sc.tl.leiden’ function in Scanpy (101) was used with the default parameter for pseudo-spot and nuclei clustering. All the clusters of pseudo-spots/nuclei were annotated by their specific gene markers. To further confirm our annotation results for each cluster, we plotted the annotated pseudo-spots/nuclei on the two-dimensional panel based on their coordinate information for each sample, and mapped the annotated pseudo-spots/nuclei on the corresponding area of the H&E sections. The orientation of each sample was confirmed by identifying the border between grey matter and adjacent white matter (WM) using the H&E stained sections coupled with the gene distribution profiles within the section (**Fig. S1A**).

#### DGE analysis

The “sc.tl.rank_genes_groups” function from the Scanpy package (v1.9.3) was utilized for DGE analysis. T-test was used, and the genes with adjusted P-value ≤ 0.05, fold-change ≥ 0.25, and mean expression across all pseudo-spots/nuclei ≥ 0.25 were considered as the DEGs.

#### Layer-layer and cell-cell communication analysis

Layer-layer and cell-cell interactions based on the expression of known LR pairs in different layer/cell types were inferred using CellChat (v2.1.0) (33). In brief, we followed the official workflow and applied the data processing functions ‘identifyOverExpressedGenes’, ‘identifyOverExpressedInteractions’. The layer and cellular communication networks were inferred by the function ‘computeCommuProb’. Function ‘netVisual_heatmap’ was used for the pairwise comparison of interactions, and the function ‘netVisual_bubble’ was used to compare the communication probabilities mediated by L-R from certain layer/cell group to other groups. All the analyses were performed with the default parameter setting.

#### Nucleus segmentation with nucleic acid staining

We followed the analysis pipeline from Stereopy (23) to capture the spatial transcriptional profiles at single-nucleus resolution. Briefly, the nucleic acid staining image of the same section for the Stereo-seq library preparation was used to project the nuclei images on the transcriptional atlas. Deep cell model (23) was used to segment the nuclei on the transcriptional atlas, and the UMI from all DNB within the corresponding segmented nuclei were aggregated per-gene and then summed to generate a nucleus by gene matrix for downstream analysis.

#### Concentric circles analysis

To understand how gene expression varies with proximity to the high stress area in PFC, we mapped the high stress focal point (identified at bin50 resolution) on a two-dimensional panel based on their coordinator for each sample, respectively, and drew three concentric circles around each high stress focal point to differentiate nuclei distances. Nuclei within a radius of 500 units of the pixel (approximately 250 µm) are categorized as level I, those between 500 and 1,500 units (approximately 250-750 µm) as level II, and those in 1,500 to 2,500 units (approximately 750-1,250 µm) as level III. Nuclei intersecting circles from multiple high stress points were assigned to the closest level (**Fig. 4D**).

#### Gene co-expression network analysis

We performed nuclei-type specific weighted gene co-expression network analysis (WGCNA) based on the nuclei from level I to III using the hdWGCNA package (76). Function ‘ConstructNetwork’ was used for co-expression network construction, and function ‘ModuleEigengenes’ and ‘ModuleConnectivity’ were used to calculate the module eigengenes and module connectivity, respectively. Function ‘GetModuleTraitCorrelation’ was used to calculate the correlation between the co-expression modules and AD levels (NA, moderate, and severe) as well as the co-expression modules and the distance to stressed neurons (from level III to I). To identify the potential transcription factors regulating the modules, the function ‘OverlapModulesMotifs’ was used to find the enriched transcription factors based on the hub genes in the module.

## Data Availability

All data produced in the present study are available upon reasonable request to the authors

## ACKNOWLEDGMENTS

The authors are grateful to STOmics Americas Ltd for technical assistance with the data generation. This work was supported by STOmics Grant Program and partially benefited from support of grants from National Institutes of Health [U19AG055373, R01AG061917, R01AG068232, P30GM145498, P20GM109036, P30AG072973].

## AUTHOR CONTRIBUTIONS

Y.G. conducted the major data analysis and wrote the main manuscript text; M.H. and R.H.S. collected the human brain samples and associated clinical information; X.Z., Y.L., and Y.P.C. performed the biological experiments; M.H., R.H.S., H.S., H.D.W, S.M.J., X.Z., X.W., A.L., L.J., D.W., Q.Z., and X.Y. provided valuable suggestions throughout the study implementation; H.S. and H.W.D. were responsible for conceiving, designing, initiating, directing, supervising, language proofreading, and securing fundings for this study. All authors participated in the discussions of the project and reviewed and/or revised the manuscript.

## DATA AVAILABILITY

The Stereo-seq data from six AD cases and six controls will be available in the GEO database after the publication of this study.

