## Supplementary figures and images for "Spatial Dissection of the Distinct Cellular Responses to Normal Aging and Alzheimer’s Disease in Human Prefrontal Cortex at Single-Nucleus Resolution"

### Supplemental figure 1

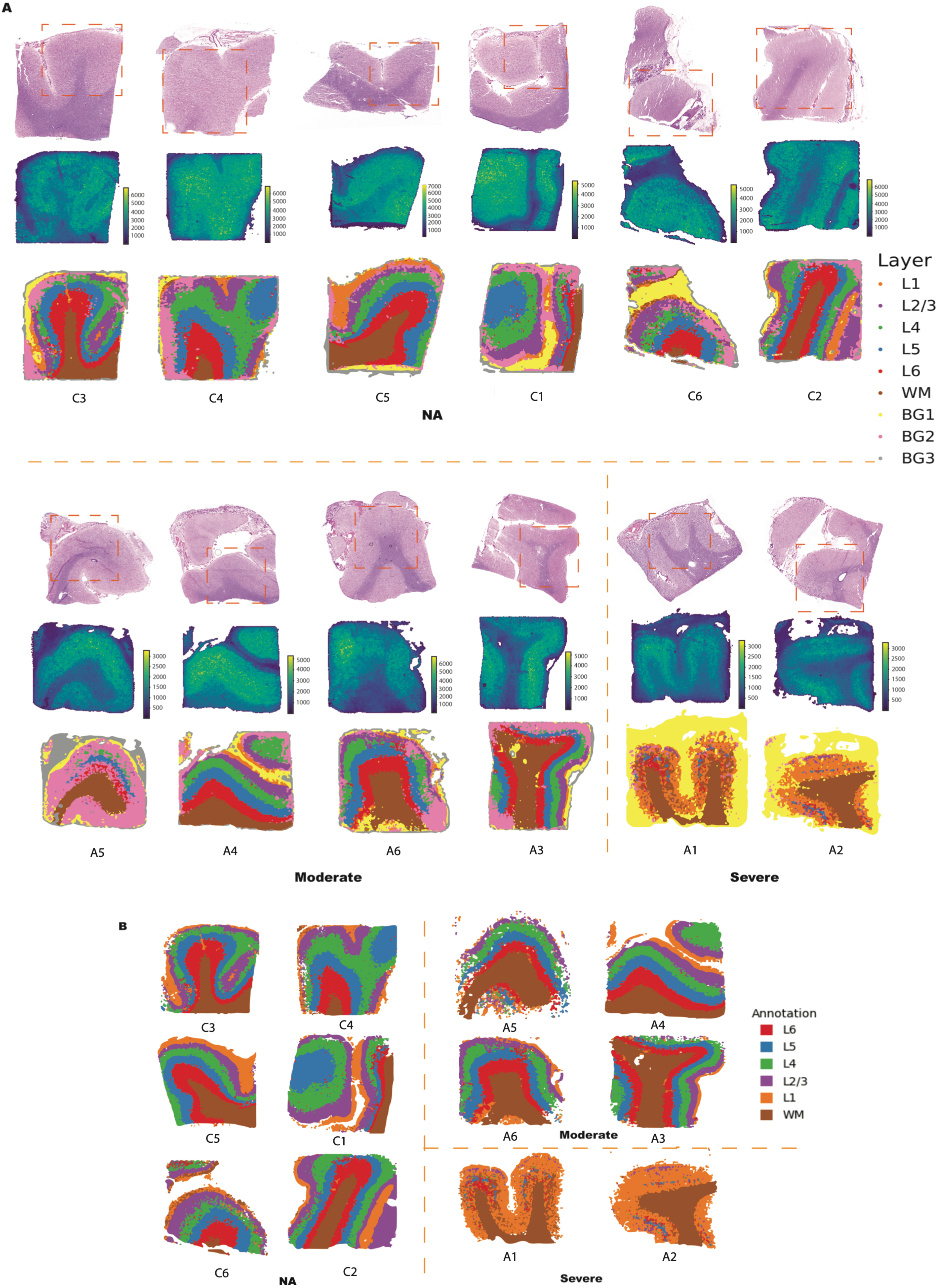

### Supplemental figure 2

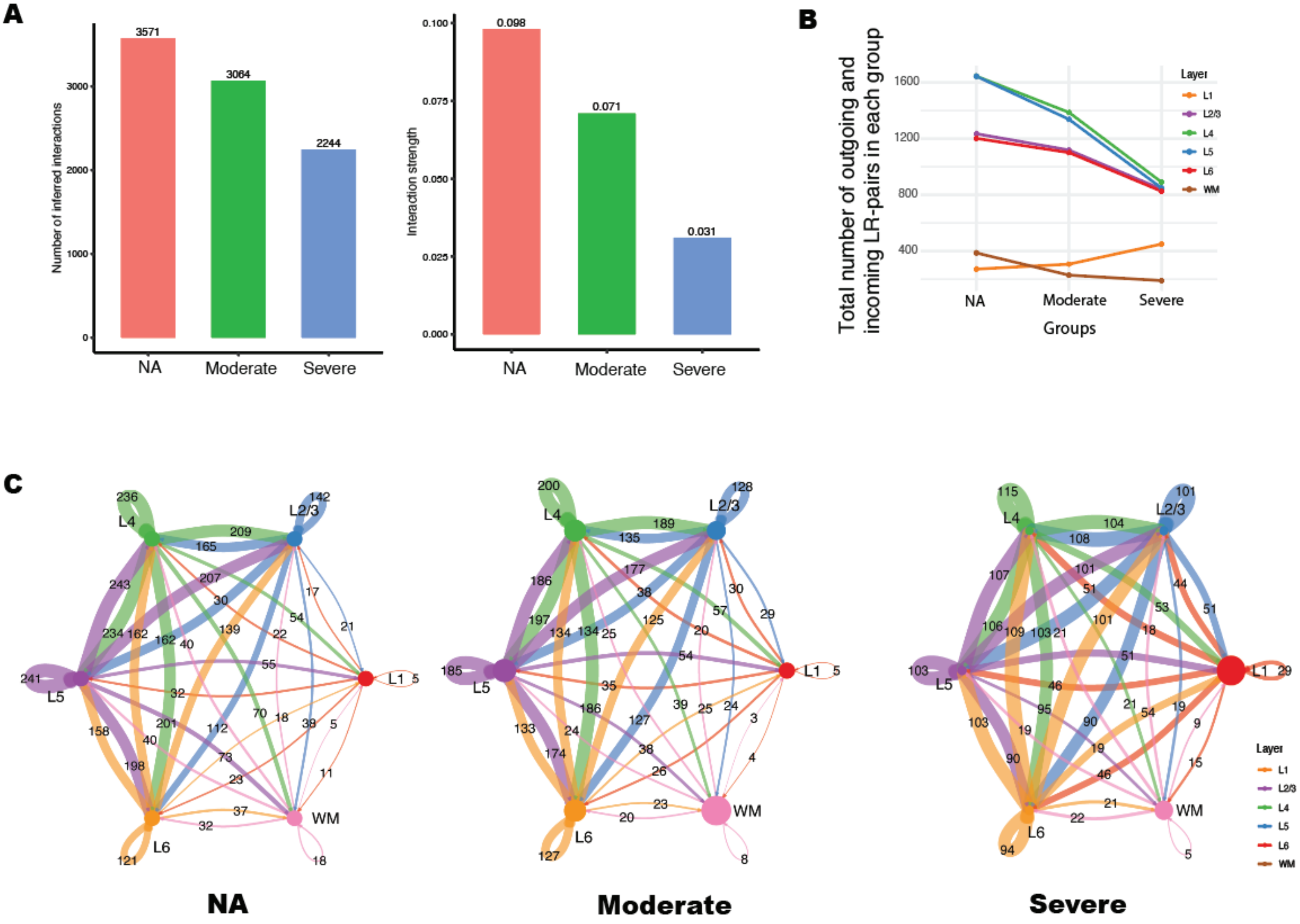

### Supplemental figure 3

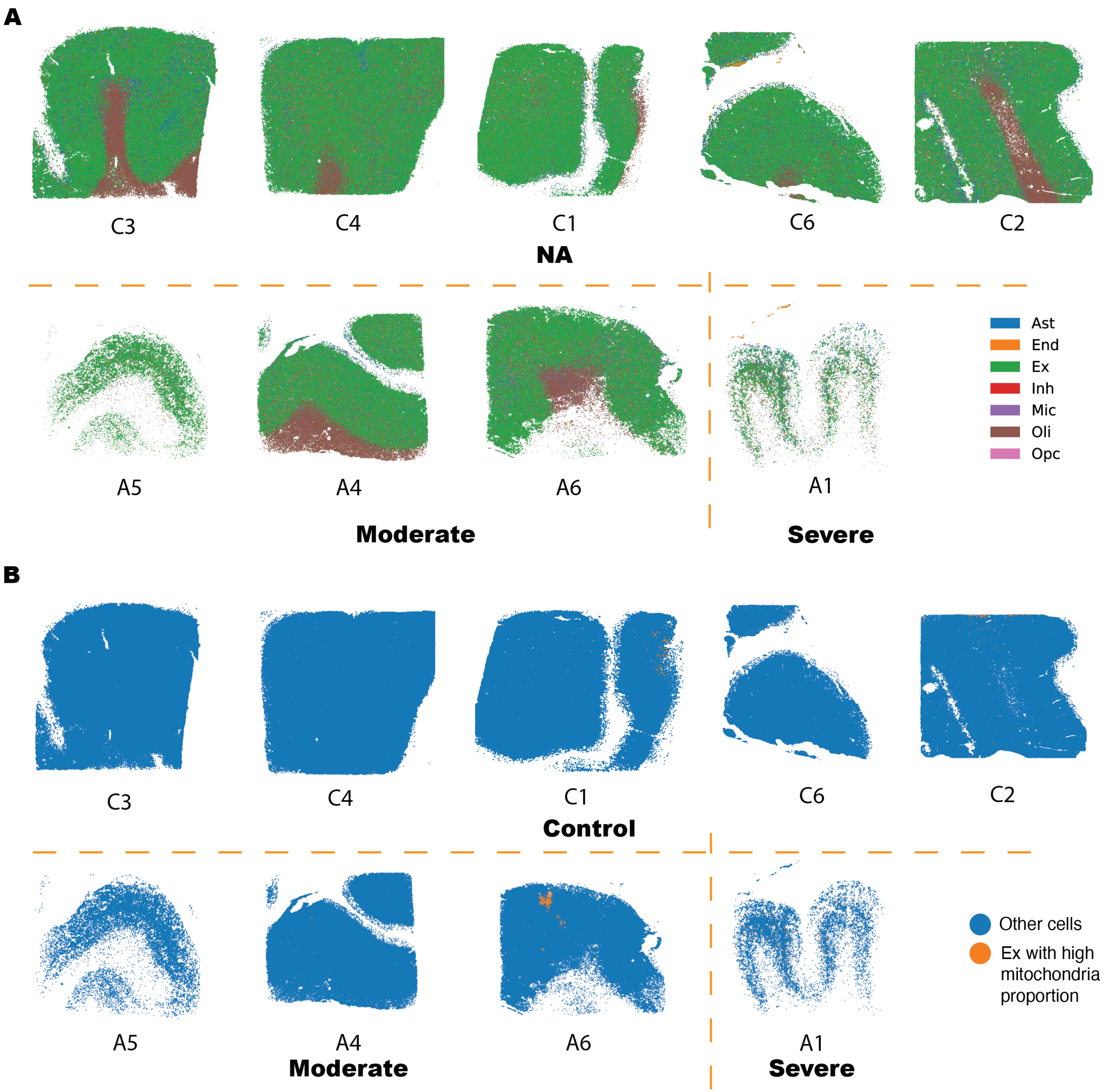

### Supplemental figure 4

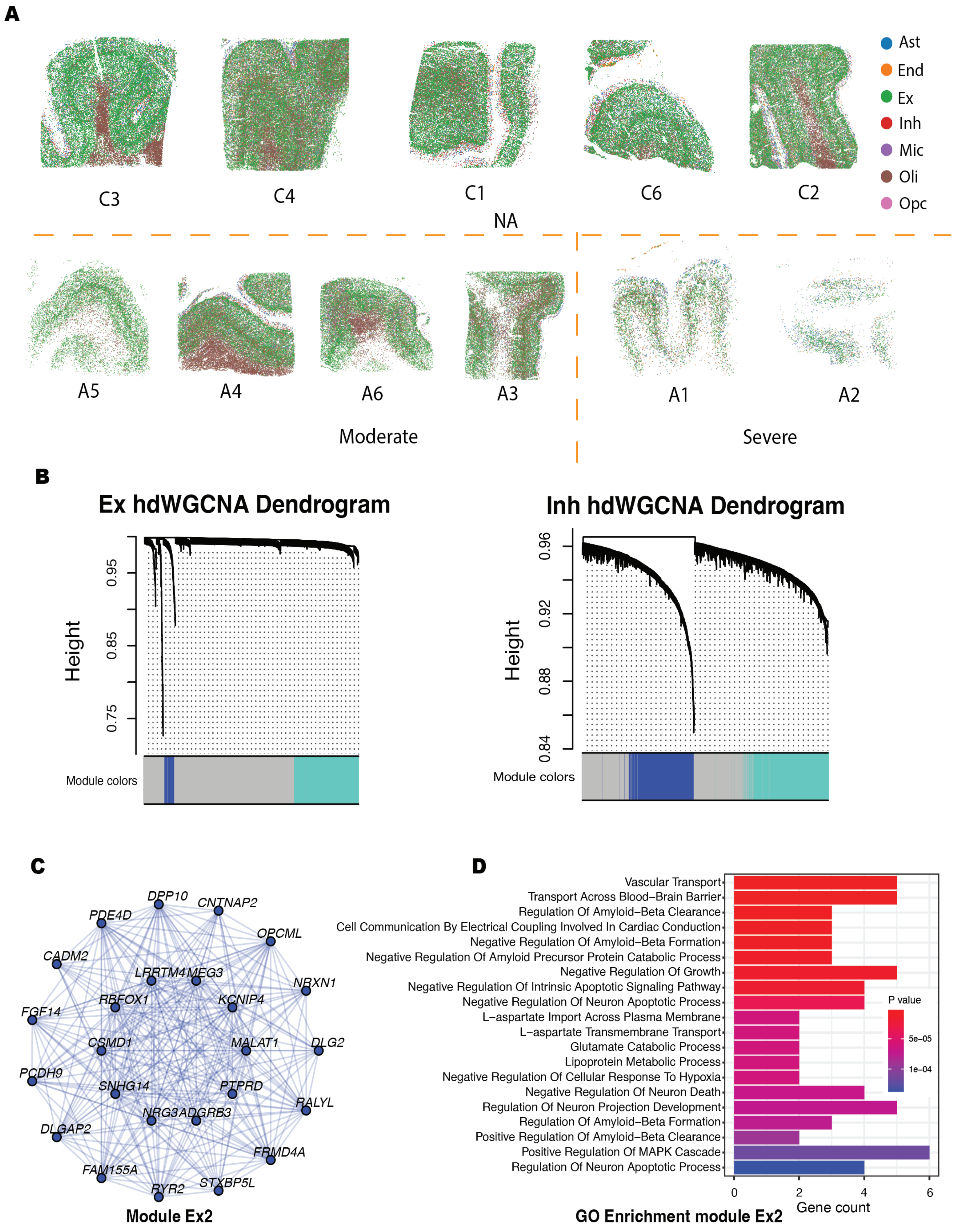
