## Supplemental figure legends and table information for "Spatial Dissection of the Distinct Cellular Responses to Normal Aging and Alzheimer’s Disease in Human Prefrontal Cortex at Single-Nucleus Resolution"

**Fig. S1: Spatial transcriptome on all samples at Bin110 resolution**

(A) The H&E staining, distribution of the total counts per pseudo-spot, and layer clustering of pseudo-spots for each sample.

(B) Scatter plot for the layer clusters after background removing.

**Fig. S2: Layer-layer interactions in the NA, moderate, and severe groups**

(A) The total number (left) and the strength (right) of the LR pairs in NA, moderate, and severe group, respectively.

(B) The total number of all ligands and receptors in each layer across NA, moderate, and severe AD groups.

(C) Networks of layer-layer interactions across six cortical layers and the WM in NA, moderate, and severe group. The colors of the dots and edges represent the specific layers and the outgoing signaling emanating from them. Number on the edge represents the number of the LR pairs.

**Fig. S3: Cell distribution on Bin50 resolution**

(A) The distribution of the annotated pseudo-spot at Bin50 resolution for NA, moderate, and severe AD groups.

(B) The distribution of highly stressed neurons all samples. Yellow spots indicate the highly stressed Ex and blue dots represent the spots annotated as other cell types (Ast, End, Inh, Mic, Oli, Opc).

**Fig. S4: Nuclei type distribution at single-nuclei resolution and gene co-expression network analysis in excitatory and inhibitory neurons**

(A) The annotated nuclei distribution across NA, moderate, and severe AD groups.

(B) The clustering dendrogram in Ex and Inh modules. Two modules (blue and turquoise) are identified in Ex and Inh, respectively.

(C) The gene co-expression modules in Ex (Ex2). The nodes represent the hub genes, and the edges between the nodes indicate the co-expression of those genes.

(D) The GO enrichment analysis on the top 50 hub genes in the Ex2 module. The length of the bar indicates the gene numbers enriched in the GO term and the color represents the adjusted P-values for enrichment analysis.

**Table S1:** Sample information for all samples

**Table S2:** Highly expressed genes in each layer and the WM across all samples

**Table S3:** DEGs of the pair-wise comparison between NA, moderate, and severe AD groups for each cortical layer and the WM

**Table S4:** Highly expressed genes in each cell types across all samples at Bin50 resolution

**Table S5:** DEGs of the comparison between nuclei in level I and III across NA, moderate, and severe AD groups

**Table S6:** The correlation of gene co-expression modules and the distance from level III to I (concentric), and AD progression (NA to severe)
